# Solvable delay model for epidemic spreading: the case of Covid-19 in Italy

**DOI:** 10.1101/2020.04.26.20080523

**Authors:** Luca Dell’Anna

## Abstract

We present a simple but realistic model for describing the diffusion of an infectious disease on a population of individuals. The dynamics is governed by a single functional delay differential equation, which, in the case of a large population, can be solved exactly, even in the presence of a time-dependent infection rate. This delay model has a higher degree of accuracy than the so-called SIR model, commonly used in epidemiology, which, instead, is formulated in terms of a set of three ordinary differential equations. We apply our model to describe the outbreak of the new virus COVID-19 in Italy, taking into account the containment measures implemented by the government in order to mitigate the spreading of the virus and the social costs for the population.

## INTRODUCTION

In a very few months a viral infection called Covid-19 (Coronavirus disease 19) originated in China, breaking through the borders of all the countries, rapidly spread all over the globalized world. Italy is one of the hardest hit countries suffering from the very dramatic consequences of this disease. The outbreak of the virus, the new coronavirus which caused the infection, seems out of our control. In the absence of a therapy and a vaccine, social distancing measures and a strict lockdown appear to be the most effective means to contain the growth of the infection. We should remind that there are places in the world where often infectious diseases, also those already defeated in the so-called more developed countries, can still cause very severe consequences among the local populations.

Even if we cannot answer the question why a virus starts spreading and which is its origin, we can still wonder how it diffuses. The aim of this work is, therefore, to provide a simple handy model for epidemic spreading, which could depend only on the couple of parameters which generally characterize an infectious disease: the infection rate and the infectiousness (or recovery) time. Both these quantities can be taken from the experience, therefore, we do not need further parameters to fit the data which could cause artificial predictions. We will show that the model we are presenting have the same, or even higher, predictive power than that of one of the most widely used technique in epidemiology, the SIR model [1–3]. This latter model requires the presence of a fictitious recovery rate related to the number of recovered persons, without considering that the new cases of recovery (and fatality) come from infected cases occurring a period of time earlier. The model we are proposing, instead, is based on the fact that the closed cases come from the infected ones after an average delay recovery time, therefore, contrary to the SIR model, formulated in terms of a set of three ordinary differential equations, it is described by just a single equation, a functional retarded differential equation, bringing predictions more under control. In this work we derive the exact analytical solution of this model in the limit of large population number, also in the presence of a time-dependent infection rate, which is the case when containment measures are implemented in order to reduce the spreading of the infection. Moreover, the definition of the so-called basic reproduction number ℛ_0_ (a parameter determining whether a infectious disease can spread or not) comes out naturally in our delay model.

We finally apply this technique to give a quantitative description of the diffusion of Covid-19 in Italy, showing the current scenario based on the actual situation and what would have happened without the containment measures. Generally it is quite difficult to give a reliable forecast on the fate of the epidemic spreading because it heavily depends on individual and social behaviors, on the effectiveness of the containment measures already implemented, or that will be taken, by the government and on the future political decisions. At the time being, even if the situation in Italy is improving, it seems that more efforts are needed in order to change course and rapidly stop the spreading of the disease. Further measures might be useful, like, for instance, i) running more diagnostic tests, at least, on all the doctors and medical workers who are in contact with many patients, ii) improving the food distribution to avoid the crowding in the food shops and to ensure subsistence goods also to those who need, iii) providing medical devices like surgical masks to all the population.

As last remark, we remind that the outbreak of Covid-19 has been declared a pandemic by the World Health Organization. Many countries are already heavily overwhelmed by this infection and by the risk for the public health, therefore, in a networked world we all have to behave and operate with an improved spirit of cooperation. The bitter lesson imparted by this tough situation is that we cannot save ourselves alone.

## THE MODEL

Let us first consider the case in which a population of individuals, subjected to an infection, is not too large or the infection is such that the recovery time for an infected person is sufficiently long. In this conditions one can expect that the epidemic diffusion is governed by the logistic equation (equivalent to the so-called SI model)

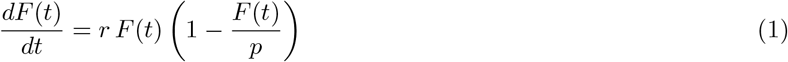

whose solution is simply given by

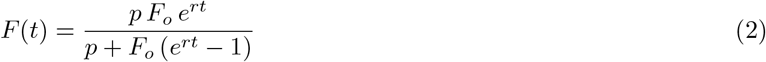

where *F*_*o*_ = *F* (*t* = 0) is the number of initial infected persons, *r* the rate of the infection, namely the number of new infections from one infected person in unit of time (the number of new infections a day), and *p* the number of individuals of the population involved. The dynamics goes on until all the population *p* is infected. This model has a poor predictable power, however, if we have enough data about the diffusion of the epidemic disease, specially in the first stage of the spreading, in order to get a rough forecast of what can happen in the near future, one could use Eq. (1) to fit the data with *F*_*o*_, *r* and *p* as free parameters.

The main issue of Eq. (1) is that it does not contain the mechanism of reduction of the spreading and the desired end of an epidemic disease. We have therefore to take into account the number of closed cases (persons who recovered or died), which do not contribute to the infection anymore. The model we are going to consider includes, therefore, the total number of infected, *F* (*t*), and the total number of recovered and deceased persons, *R*(*t*), so that Eq. (2) becomes

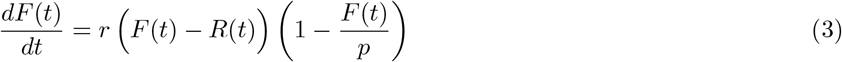

In principle also *R*(*t*) can follow another dynamical equation, however, generally, there is an average time of recovering *δt* so that the total number of cases at some time *t* becomes closed cases at later time *t* + *δt*, namely

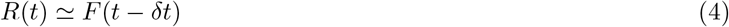

This seems to be the case also for the new coronavirus spreading, by looking at some reported data for Covid-19 in Italy, shown in Fig. 1 (see also Ref. [4]). Eq. (4) allows us to write Eq. (3) in terms of only the function *F* (*t*). Notice that, as we will discuss later, Eq. (3) can be derived also from the SIR model. If we consider the case where the population *p* is very large, as long as *F* (*t*) *≪ p*, we can neglect the logistic term, 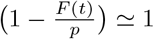, so to have

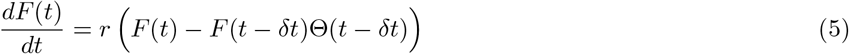

where Θ is the Heaviside theta function. Eq. (5) is a functional retarded differential equation.

**FIG. 1:**
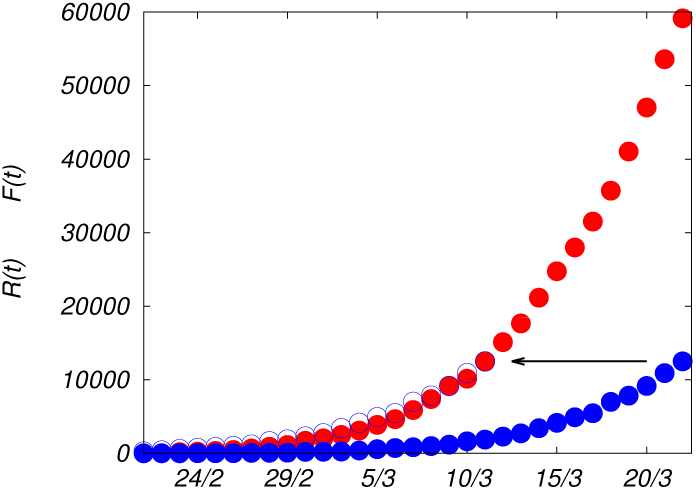
Total number of confirmed cases of Covid-19 in Italy, *F* (*t*) (red dots), reported in Ref. [5], since 21th February to 22th March 2020, compared with the closed cases, *R*(*t*) (blue dots), in the same period of time. If the numbers of closed cases are shifted in time by *δt* ≃ 11 days (blue circles) they fairly overlap with the total numbers of cases.

## EXACT SOLUTION

Writing the time *t* as *t* = *n δt* + *t* ′, where 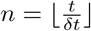 is the integer part of *t/δt*, the solution of Eq. (5) is given by

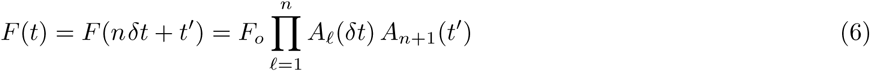

where the functions *A*_*ℓ*_ fulfill the following iterative equation

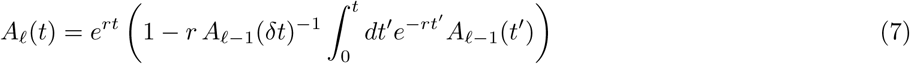

with *A*_0_(*t*) = 0 for any *t < δt* and *A*_0_(*δt*) = 1, so that, for *ℓ* = 1, we recover *A*_1_(*t*) = *e*^*rt*^. The full exact solution is, therefore, obtained by solving a cascade of *n* local integrals. The proof of Eqs. (6) and (7) is given in Appendix A.

At time *t* = *n δt*, from Eq. (7), performing the chain of integrals, and putting the results in Eq. (6), we get the following exact result,

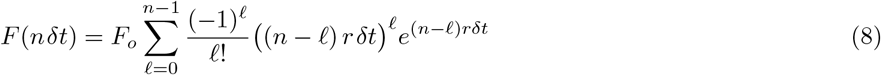

For instance, for *n* = 1 and *n* = 2, namely up to twice the infectiousness period, the total number of cases is simply

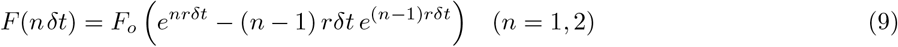

Surprisingly we find that Eq. (8) depends only on (*r δt*), which is called basic reproduction number ℛ_0_ (see below). It is easy to check from Eq. (8) that, while for large ℛ_0_ = *rδt, F* (*nδt*) is dominated by an exponential behavior, for ℛ_0_ = 1, *F* (*nδt*) becomes linear in *n*. From Eq. (6) and Eq. (7), we can notice that the function *F* (*t*) depends on its past, therefore, it seems governed by a non-Makovian dynamics. Once we have the total number of infections *F* (*t*), we can also calculate the number of persons who are still infected, at a given time *t*, which is defined by

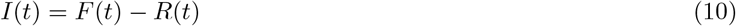

In our model *I*(*t*) = *F* (*t*) − *F* (*t −δt*)Θ(*t − δt*), so that Eq. (5), in terms of this quantity is simply 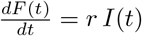. Before to proceed, a comment on the comparison with another model is in order. The so-called SIR model is one of the most used techniques for describing the spreading in time of an infection disease. According to this model the population is divided into three parts represented by the number of susceptible *S*(*t*), infected *I*(*t*) and recovered *R*(*t*) individuals which vary over time (see Appendix B). This model, is almost equivalent to our simpler model, Eq. (3)-(4). However a criticism which can be raised against the SIR model is related to the fact that, being formulated in terms of ordinary differential equations, the model requires the presence of an effective recovery (and fatality) rate which might not correspond to the actual rate since the new cases of recovery (and fatality) come from infected cases occurring a few days earlier. For that reason, instead of writing the problem in terms of ordinary differential equations one has to do it in terms of functional differential equations.

### Basic reproduction number

Let us consider Eq. (5), for *t > δt*, in the following form

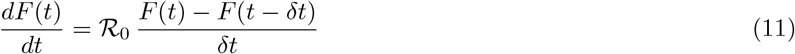

where we introduce and naturally identify ℛ_0_ as the so-called basic reproduction number

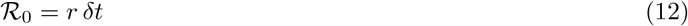

which is a widely used parameter for predicting whether the infectious disease will spread into a population or turns off, and represents the average number of cases originated by a single infectious case during the infectiousness period. Eq. (11) implies that the first derivative of *F* (*t*) is equal to its increment in a time interval *δt*, divided by *δt*, namely *F* (*t*) is linear in *t* if the rate is equal to the critical value *r* = *δt*^*−*1^ (ℛ_0_ = 1). For *r > δt*^*−*1^ (ℛ_0_ *>* 1), the function *F* (*t*) increases more than linearly, while for *r < δt*^*−*1^ (ℛ_0_ *<* 1), *F* (*t*) goes slower than linearly. If we let *r* vary in time, when *r* = *δt*^*−*1^ (ℛ_0_ = 1) the function *F* (*t*) has an inflection point, where it changes from being concave to convex or vice versa. Making a comparison with the SIR model, where ℛ_0_ = *r/β*, one can identify *β*, the fictitious recovery rate (see Appendix B) with the inverse of the recovery time *β ∼* 1*/δt*.

### Time-dependent infection rate: exact solution

Let us now consider the possibility of having a time-dependent infection rate *r*(*t*) in the dynamical equation for the total number of infected persons

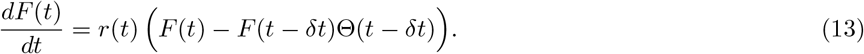

Also in this more general case the exact solution, valid for any profile of *r*(*t*), can be written in the same form of Eq. (6), namely, 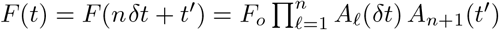, where now the functions *A_ℓ_* are given by

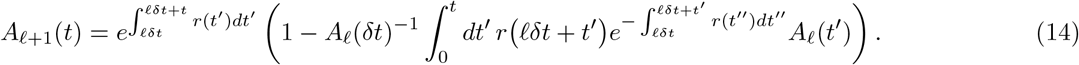

For instance, 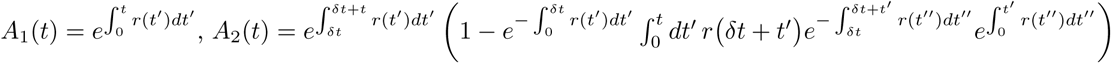, and so on. For constant *r*, Eq. (14) reduces to Eq. (7). See Appendix A for more details about the derivation. The solution *F* (*t*) has therefore to fulfill the following recursive equation, after splitting the time in *n* intervals *δt* with the residual time *t* ′

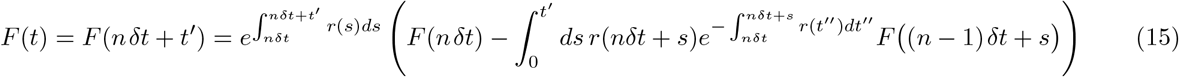

This general result implies that if we knew the time dependence of the infection rate or if we could tailor its evolution by, for instance, containment measures, we can know the exact analytical expression of *F* (*t*), the total number of infected persons, as a function of time, as long as *F* (*t*) is much smaller than *p*.

## COVID-19 IN ITALY

Let us consider the delay model in its general form Eqs. (3)-(4), where the infection rate *r* varies in time

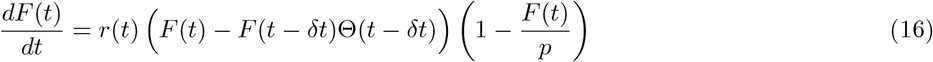

as the effect of some containment measures taken in order to reduce the impact of an infection on the population. As an example, let us suppose that *r*(*t*) is modified by social distancing measures, lockdown and the shutdown of many work activities, as in it is happening in Italy (and in many other countries) to mitigate and reduce the spreading of the new coronavirus, Covid-19, after two main decrees imposed by the Italian Prime Minister ordering the lockdown of the whole national territory, taken on March 11-th (lockdown and shutdown of many stores) and March 22-th 2020 (shutdown of many factories and strengthening of social distancing measures), after some other measures taken right before for local regions (e.g. the decree of March 8-th for the lockdown of Lombardy and other areas). As a result, we can imagine that *r*(*t*) decreases smoothly after those dates taking into account the adaptation time for the individuals to the new social behaviors and the period needed to complete the last activities before the blockade of the factories. Let us suppose, therefore, that *r*(*t*) can change in time according to a smooth step function as in Eq. (17),

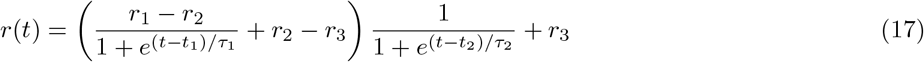

where *t*_1_ and *t*_2_ are the times where the steps are located, *τ*_1_ and *τ*_2_ make the function to be smooth, *r*_1_ is the initial observed infection rate which causes the starting exponential growth of the epidemic disease, *r*_2_ the intermediate rate, which fits with the data, supposed to be reached after the first decree of lockdown, and *r*_3_ the supposed asymptotic infection rate after the second decree of lockdown. Fixing the average of recovery and fatality time *δt*, the basic reproduction number is also a function of time according to ℛ_0_ = *r*(*t*)*δt*, with a profile shown in Fig. 2.

**FIG. 2:**
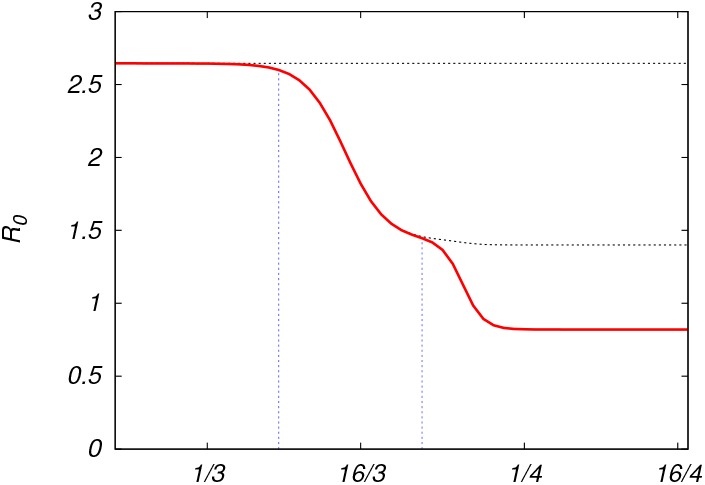
Basic reproduction number ℛ_0_ = *r*(*t*)*δt*, as a function of time, based on the profile for the infection rate described by Eq. (17). We take *δt* about 11 or 12 days [4], *t*_1_ between 13th and 14th March 2020, *t*_2_ on 26th March, *τ*_1_ ∼ 2 days, *τ*_2_ ∼ 1 day. The starting value is ℛ_0_ = *r*_1_*δt* ≃ 2.65 and the intermediate value is ℛ_0_ = *r*_2_*δt* ≃ 1.45, and a current value, ℛ_0_ = *r*_3_*δt* ≃ 0.85. The vertical dotted blue lines point the dates of the main laws for the containment measures (8th-11th March and 22th March 2020). The gray dotted lines correspond to ℛ_0_ in the absence of the first and the second containment measures.

Solving Eq. (16), or, analogously, using the recursive relation in Eq. (15), with the time-dependent rate *r*(*t*) given by Eq. (17), fixed by the parameters producing the profile depicted in Fig. 2, we obtain the solution *F* (*t*) which slowly goes to saturation over time, in perfect agreement with the data for the total number of confirmed infected cases, as shown by Fig. 3, where the blue line is the expected curve, while the red points are the official data. The dotted gray lines in Fig. 3 represent *F* (*t*) if the containment measures had not been taken. As one can see from Figs. 2-3, only when ℛ_0_ becomes smaller than 1, the curve flattens allowing for a stop of the epidemic spreading, avoiding that a large part of the population gets infected. For ℛ_0_ ≃1, *F* (*t*) would increase linearly, and *I*(*t*) would become almost constant, meaning that the number of new infections would be equal to the number of closed cases. A reliable forecast has to take into account the fact that the official data of infectious cases are made by counting mostly the symptomatic cases, probably discarding other infectious cases which could transfer the virus even without or with mild symptoms. Moreover, the data of both the total number of infected persons and that of the recovered ones could be affected by the procedure, the realization times and the number of the diagnostic tests. However, since our model relies on the infectiousness time, it does not need a fitting of the data for recovered persons which may be affected by systematic errors. This uncertainty on the data for closed cases would compromise the result for the SIR model. On the contrary, our theoretical prediction based on the delay model agrees fairly well with the data-set for total infected cases, as shown in Fig. 3.

**FIG. 3:**
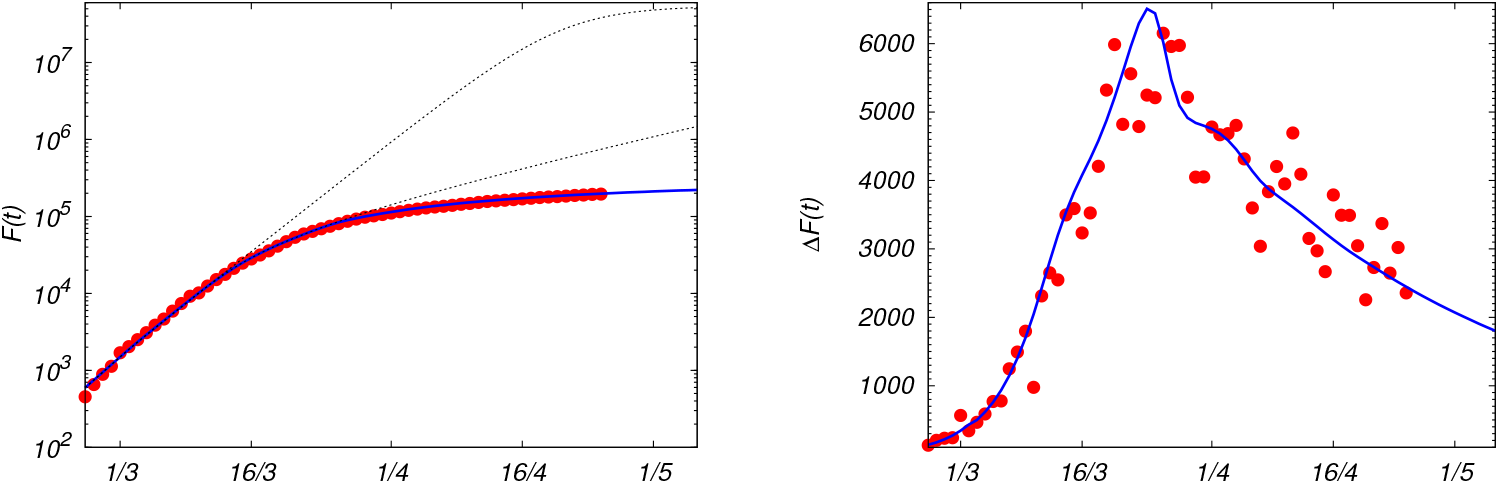
(Left) Total number of infected persons over time, *F* (*t*) (red points), from official data for Covid-19 in Italy [5], where *p* = 6 · 10^7^, up to 14th April. The blue line is the theoretical prediction *F* (*t*) as solution of Eq. (16), with initial conditions, fixed at *t* = 0 the 21th February, *F*_*o*_ ≃150, and *r* = *r*_1_ ≃ 0.23 (ℛ_0_ ≃ 2.65), and using the profile for the infection rate given by Eq. (17), with the parameters reported in Fig. 2. The gray dotted lines are the expected curves for *F* (*t*) if the first and the second containment measures (8-11th March and 22th March) had not been taken. (Right) Daily number of infected persons, Δ*F* (*t*), compared with the theoretical 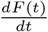 from the solution of Eq. (16).

As a final remark we remind that most of the confirmed infected cases in Italy are counted after the appearance of the symptoms and the persons who exhibit severe ones are mostly hospitalized, and then counted as infected persons. Some of them, unfortunately, die approximately 4 days after (therefore after approximately 9 days form the onset of the first symptoms, as reported by the Istituto Superiore di Sanitá, www.iss.it). We observe that, splitting the closed cases between real recovered persons, *R*_*R*_, and dead persons, *R*_*D*_,

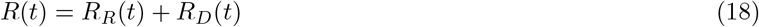

and since the confirmation of recovery needs extra diagnostic tests which are still not widely performed, the most reliable data is that of dead persons *R*_*D*_(*t*), which is found to be linked to the total number of confirmed infected cases, *F* (*t*), in the following way

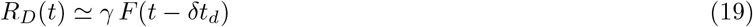

with 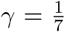 and a delay time of *δt*_*d*_ = 4 days, as show in Fig. 4. The number of victims follows the number of total confirmed cases and it is equal to 1*/*7 of its value four days before.

**FIG. 4:**
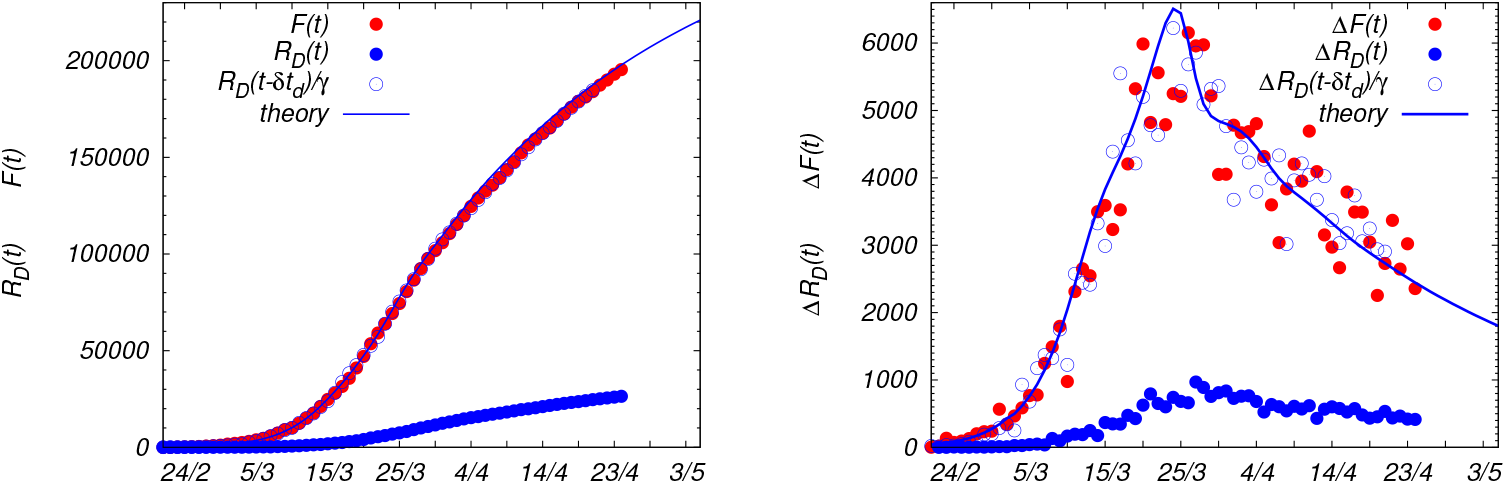
(Left) Total number of confirmed cases of Covid-19 in Italy, *F* (*t*) (red dots), reported in Ref. [5], up to 25th April 2020, compared with the decesed cases, *R*_*D*_ (*t*) (blue dots), in the same period of time. If the numbers of died persons are shifted in time by *δt*_*d*_ ≃ 4 days and rescaled by *γ*^*−*1^ = 7 (blue circles) they perfectly overlap with the total numbers of cases. The blue solid line is theoretical prediction for *F* (*t*), solution of Eq. (16). (Right) Daily number of infected persons, Δ*F* (*t*) (red dots), and daily number of victims Δ*R*_*D*_ (*t*) (blue dots). The blue circles are the daily number of victims after rescaling according to Eq. (19), with *δt*_*d*_= 4 days and *γ* = 1*/*7. The solid blue line is 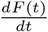 from the solution of Eq. (16).

The fatality of the sick persons, those who exhibit some symptoms, is therefore quite high, 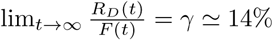.

## CONCLUSIONS

We present a simple but realistic model for describing epidemic spreading, based on the fact that the closed cases come from infected ones at an early time. This observation allows us to formulate the problem in terms of a single functional differential equation depending on two well defined clinically relevant parameters: the infection rate and the infectiousness time. We provide the exact analytical solution for such an equation, in the limit of large population number, finding that it depends exclusively on the basic reproduction number ℛ_0_ = *rδt*, see Eq. (8). We derive the analytic solution also in the presence of a generic time-dependent infection rate. We apply our model to the case of the spreading of Covid-19 in Italy, allowing the infection rate to vary in time, as a result of some containment measures implemented by the government in order to mitigate the consequences of the infection on the population. We find perfect agreement between the official data and the expected theoretical results. In general terms, the basic reproduction number should be suppressed well below 1 in order to rapidly recover the initial condition. By a rough estimation, in order to have a decline of the infection as fast as its growth, containment measures or possible therapies should be so effective to reduce the basic reproduction number and reach the final value *R*_0*f*_ such that 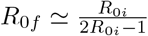, starting from an initial value *R*_0 *i*_. In the case of Covid-19 in Italy, the initial value for the basic reproduction number was *R*_0*i*_ *≃* 2.6, while the current one seems to settle at *R*_0*f*_ *≃* 0.8, implying a rather slow decline of the infection. Finally we discussed the fatality rate, showing that the number of victims is exactly a fraction of the total number of cases few days before.

## Data Availability

The data used in the manuscript are taken from https://lab.gedidigital.it/gedi-visual/2020/coronavirus-i-contagi-in-italia/

## Appendix A: Solution of the retarded differential equation

For *t ≤ δt*, the solution of Eq. (5) is *F* (*t*) = *F*_*o*_*e*^*rt*^. Let us consider *t* = *δt* + *dt* with infinitesimal *dt*, from Eq. (5)

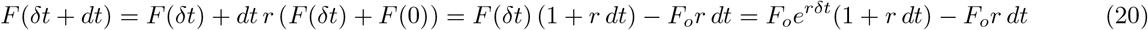

Using this result we can calculate

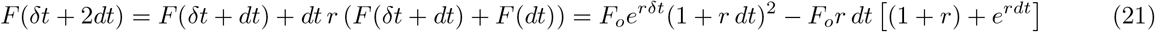

Analogously, from that, we can proceed calculating

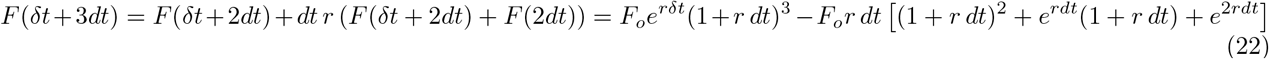

and going on by adding infinitesimal time steps, we find iteratively that

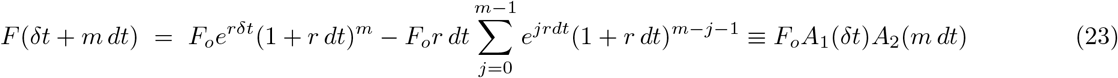

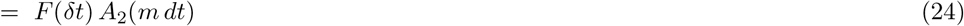

with *A*_1_(*δt*) = *e*^*rδt*^ and defining

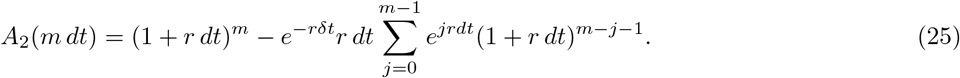

In particular, for *m dt* = *δt*, we have an expression for *F* (2*δt*) in terms of the function at early time, *F* (2*δt*) = *F* (*δt*)*A*_2_(*δt*). We can now start again with the iteration

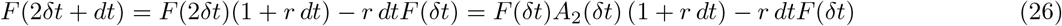

One can proceed in the same way as before getting

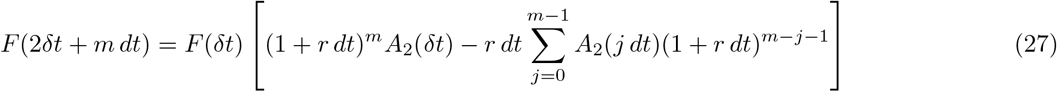

which can be written as

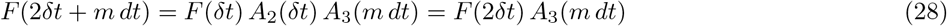

where

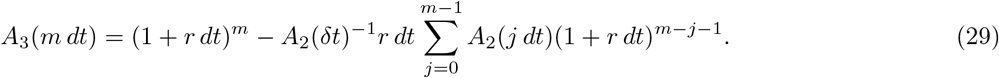

We can notice that at any step *δt* we can perform the same calculation since we can factorize the function *F* as

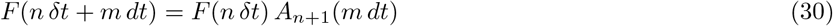

where, therefore, 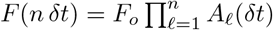 and

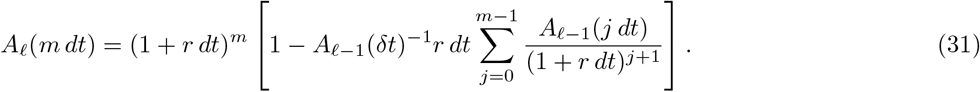

In the continuum limit, *dt →* 0 and *m → ∞*, keeping finite the time interval *m dt* = *t*, reminding that

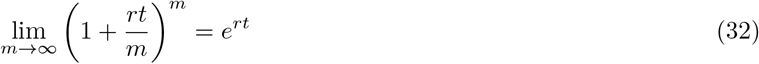

we finally obtain the result reported Eq. (7).

In the presence of time dependent infection rate, splitting again the time in *n* intervals *δt* and the residual time in *m* infinitesimal intervals *dt*, we define

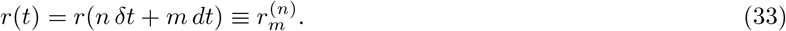

Proceeding iteratively as done for the constant rate case, but now taking trace of the different values of *r*,

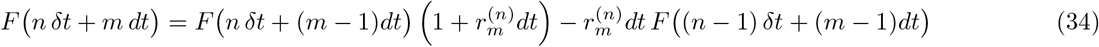

after several steps, similar to those done previously, we find that Eq. (31) can be generalized in the following way

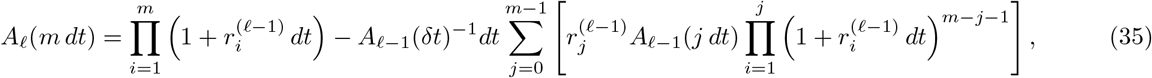

whose continuum limit is given in Eq. (14).

## Appendix B: Comparison with the SIR model

The most commonly used model for epidemic spreading is the so-called SIR model, which describes the dynamics of the number of susceptible, *S*(*t*), infected, *I*(*t*) and recovered, *R*(*t*) persons, according to the following differential equations

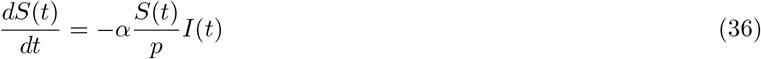

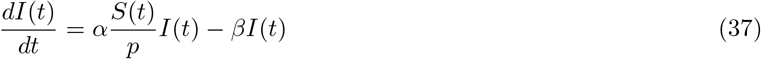

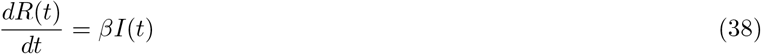

with generally the initial condition *S*(0) ≃ *p*. The free parameters *α*, the infection rate and *β*, the recovery rate, can be fixed by fitting the data sets. Defining

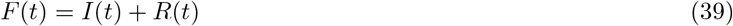

and summing Eqs. (37)-(38) we get

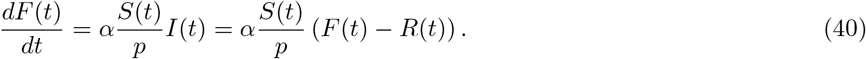

Summing the three Eqs. (37)-(38) one gets, for any time *t*,

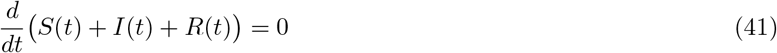

namely that, the sum of the three functions is constant and equal to the population *p* for any *t*, since at the beginning *S*(0) + *I*(0) = *p*, therefore,

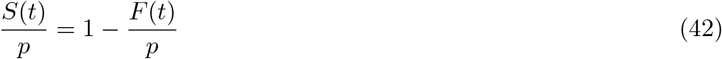

which is nothing but the logistic term so that Eq. (40) is exactly equal to Eq. (3), where *α* = *r*. This implies that one equation among Eqs. (36)-(38) is redundant, therefore, instead of considering three equations one can take just two. For instance, we can choose to express the time evolution in terms of *F* (*t*) and *R*(*t*),

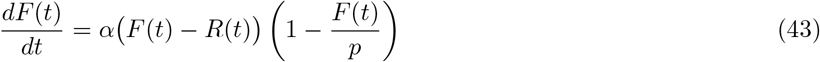

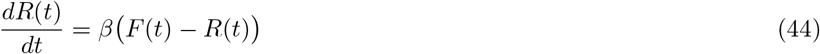

In particular, as long as *F* (*t*) *≪ p* so that (1 *− F* (*t*)*/p*) *≈* 1, from Eqs. (43) and (44), we have

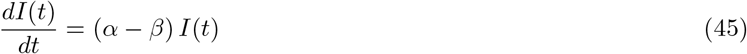

meaning that only if *α* is smaller than *β, α < β*, the infection shuts down. One can introduce the so-called basic reproduction number ℛ_0_ which predicts whether the infectious disease will spread into a population or die out, and represents the average number of cases originated by a single infectious case in a totally susceptible population during the infectiousness period. This quantity is defined as

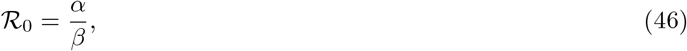

and, from Eq. (45), one can see that only for ℛ_0_ *<* 1 the infection turns off.

We notice that Eq. (43) is equal to Eq. (3). The difference between the delay model, described by Eqs. (3)-(4), and the SIR model, described by Eqs. (43)-(44), is that, in our model, the number of closed cases *R*(*t*) is locked to be equal to the total cases at a former time *F* (*t −δt*), an average recovery period *δt* before. In other words, in the delay model Eq. (44) is substituted by Eq. (4), which is equivalent to Eq. (44) with *β* = *α* but with a different initial condition, *R*(*δt*) = *F* (0). The delay time allows us to define the basic reproduction number more naturally as

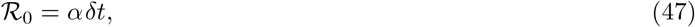

the number of new infections during the infectiousness time *δt*. On the contrary, in the SIR model one has to artificially adjust the rate *β* to take into account the right initial condition.

